# Impact of sequential adjustment on the association between metabolic syndrome and stroke: A population-based study in Korea

**DOI:** 10.64898/2026.06.27.26356735

**Authors:** Seung Han Chae, Il Young Moon, Chung Hwi Yi

## Abstract

**Background:** Stroke is among the foremost contributors to mortality and lasting disability and continues to place a heavy clinical and societal burden worldwide. Although metabolic syndrome (MetS) is recognized as a contributor to stroke risk, insufficient attention has been paid to how this relationship behaves when potential confounders are entered into the model in a stepwise manner.

**Objectives:** This study aimed to characterize the relationship between MetS and stroke prevalence using a series of sequentially adjusted models built from the Korea National Health and Nutrition Examination Survey (KNHANES). We explored whether self-rated health is a useful functional indicator for stratifying stroke risk.

**Methods:** Of the 22,559 KNHANES VIII respondents, 12,536 participants aged ≥19 years and with information required to classify physician-diagnosed stroke (DI3_dg) or define MetS were included in the final analysis. Associations were estimated using complex-sample logistic regression under a sequential adjustment scheme: Model 1 (unadjusted), Model 2 (adjusted for age and sex), Model 3 (further adjusted for educational level and family history of stroke), and Model 4 (additionally incorporating economic activity status and self-rated health).

**Results:** MetS was associated with an increased risk of stroke in all models: Model 1 (odds ratio [OR] 3.372, 95% confidence interval [CI] 2.443–4.654), Model 2 (OR 1.956, 95% CI 1.396–2.740), Model 3 (OR 1.813, 95% CI 1.292–2.546), and Model 4 (OR 1.636, 95% CI 1.153–2.321). A graded pattern was noted concerning self-rated health, with progressively poorer perceived health corresponding to higher odds of stroke, and the “very poor” category showed substantially elevated odds (OR 9.836 in Model 4).

**Conclusions:** MetS was independently associated with stroke prevalence even after sequential adjustment. Self-rated health appears to capture both metabolic burden and broader functional health aspects.

## Introduction

Stroke ranks among the leading causes of death and long-term disability and is associated with substantial clinical and economic burdens [1]. The worldwide stroke burden continues to increase owing to widespread population aging, with increasing numbers of individuals living with chronic stroke-related disabilities [2]. In South Korea, the combined effects of rapid population aging and recent advances in the acute management of stroke have shifted the burden of disease away from early mortality toward long-term functional limitations and sustained care needs [3]. In this context, identifying modifiable contributors to stroke risk has become essential for primary prevention and stratification of stroke risk at the population level [3,4].

Metabolic syndrome (MetS) denotes a clustering of cardiometabolic disturbances, such as central adiposity, raised blood pressure, dyslipidemia, and disturbed glucose handling, and is widely regarded as a substantial contributor to cardiovascular disease and stroke [5,6]. Patients with MetS have a higher likelihood of developing incident stroke than those without [6,7]. Estimates of the extent of this relationship differ considerably between studies; however, this raises questions regarding whether these observations actually reflect differences in the demographic makeup, socioeconomic circumstances, and overall health profiles of the populations examined [8,9].

The risk of stroke is influenced by structural health determinants, such as age, socioeconomic status, and access to healthcare [10,11]. These factors may confound or contextualize the association between metabolic disturbances and cerebrovascular outcomes [11]. Thus, some of the apparent associations between MetS and stroke may stem from shared demographic and socioeconomic characteristics rather than a strictly biological connection between metabolic dysregulation and cerebral vascular disease [10].

Subjective health status, commonly captured as self-rated health (SRH), has gained increasing recognition as a meaningful predictor of illness and death [12]. SRH expresses an individual’s overall, integrated sense of their health and draws on physical, psychological, and social dimensions that objective clinical measures may not fully reflect [13]. Poor SRH is associated with a higher risk of cardiovascular events and mortality, suggesting that it may capture a broader form of vulnerability beyond that captured by conventional biomedical risk markers [14].

Although the association between MetS and stroke is well documented, few population-based investigations have directly quantified the extent to which it is attributable to demographic, socioeconomic, and subjective health-related characteristics [15,16]. This knowledge gap is substantial, as it remains unclear whether the observed relationship is driven mainly by metabolic pathology or the wider health-related context within a given population. Sequential adjustment offers a practical means of addressing this gap, as it shows how an exposure–outcome estimate shifts when sets of potential confounders are introduced individually [17,18]. Examining this progressive change may help establish whether the MetS–stroke relationship primarily reflects metabolic mechanisms or is contextual [15,18]. Thus, this study used nationally representative data from the Korea National Health and Nutrition Examination Survey (KNHANES) to examine the relationship between MetS and physician-diagnosed stroke among Korean adults. We aimed to evaluate how the estimated association changed across sequential models that were adjusted for demographic characteristics, socioeconomic factors, and SRH to clarify the contextual structure underlying the observed MetS–stroke relationship.

## Methods

### Study design and participants

We analyzed data from the Eighth KNHANES (KNHANES VIII), which was conducted in 2019–2021, using a cross-sectional design. This nationally representative survey, administered by the Korea Disease Control and Prevention Agency, uses a stratified, multistage, probability-based cluster sampling scheme to represent the non-institutionalized population of South Korea. Data were gathered through health interviews, standardized physical examinations, and laboratory testing performed by trained survey staff.

Figure 1 outlines the selection of the study participants. Of the 22,559 total KNHANES VIII respondents, only those aged ≥19 years (n = 18,691) met the eligibility criteria for this study. We excluded those who answered “don’t know” or had missing responses for the physician-diagnosed stroke item (DI3_dg) (n = 1,976), those missing any of the five components required to define MetS (n = 2,919), and those with incomplete information concerning the covariates of interest (parental history of stroke, self-rated health, or economic activity status; n = 1,260). The final analytical sample comprised 12,536 individuals.

**Fig. 1.**
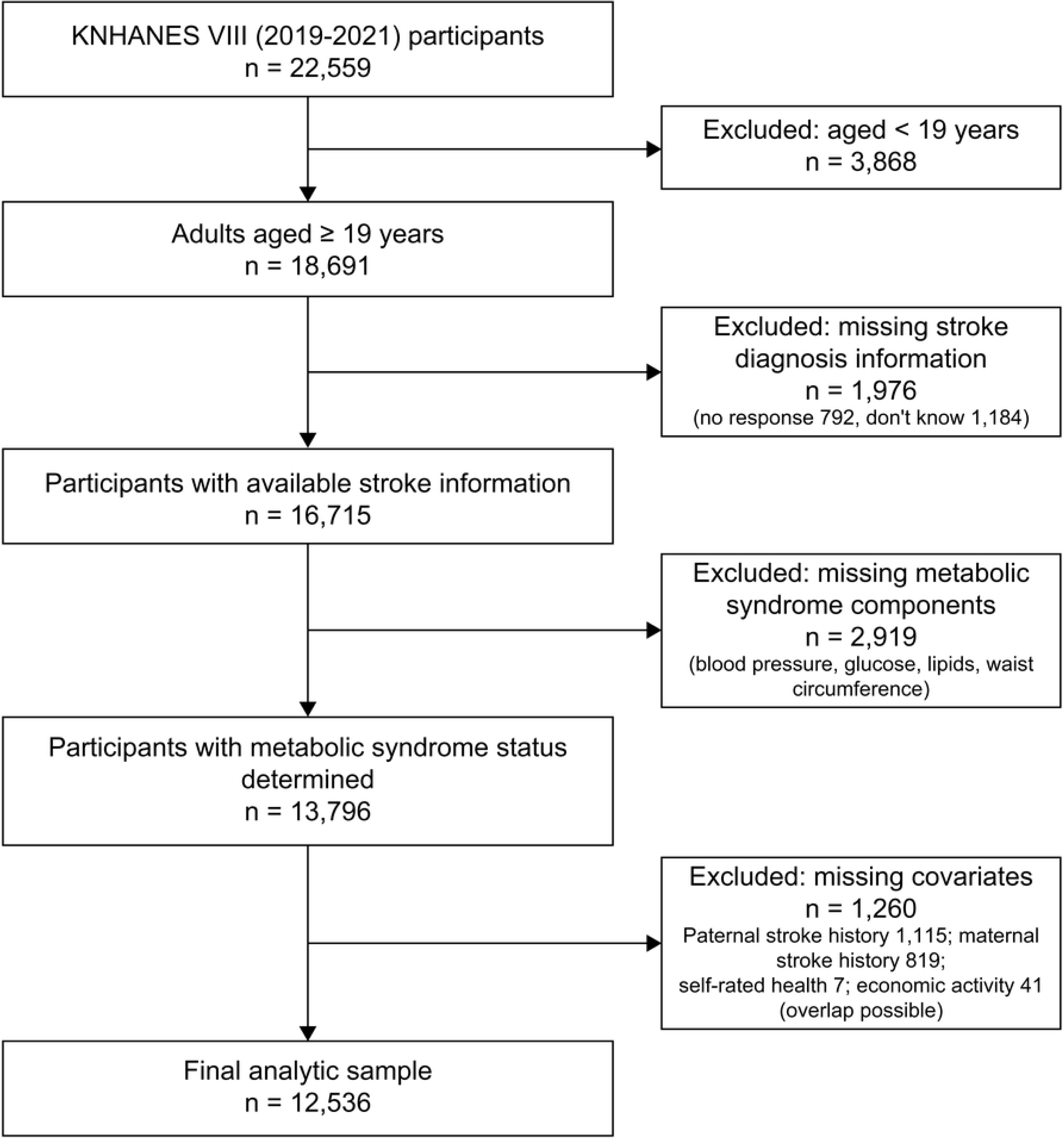
Flow Diagram of Participant Selection. KNHANES = Korea National Health and Nutrition Examination Survey. The sum of individual exclusion reasons in the final step exceeds the total excluded (n = 1,260) because some participants had missing values on more than one covariate.

To respect the complex survey design and preserve the national representativeness of the data, every analysis applied the integrated 3-year sampling weight (the combined health interview and examination weight [wt_itvex] divided by three) along with the stratification variable (kstrata) and the primary sampling units (psu).

### Measures

#### Stroke

Stroke data were ascertained from the KNHANES health interview items regarding physician-diagnosed stroke (DI3_dg). Respondents who answered “Yes” to the question “Have you ever been diagnosed with stroke by a physician?” were recorded as having a history of stroke, whereas those answering “No” were considered stroke-free. Responses of “I don’t know” (DI3_dg = 9) and non-responses were dropped. Stroke was considered a binary outcome (0 = no history; 1 = physician-diagnosed stroke).

#### MetS

MetS was defined using the criteria of the Korean Society for the Study of Obesity, which have been tailored to the characteristics of Asian populations. Thus, participants were classified as having MetS when at least three of the following five criteria were satisfied:

- Abdominal obesity: waist circumference ≥ 90 cm (men) or ≥ 85 cm (women);
- Elevated blood pressure: systolic blood pressure ≥ 130 mmHg, diastolic blood pressure ≥ 85 mmHg, or current use of antihypertensive medication;
- Elevated fasting glucose: fasting plasma level ≥ 100 mg/dL or currently receiving treatment for diabetes;
- Hypertriglyceridemia: triglycerides ≥ 150 mg/dL;
- Reduced high-density lipoprotein cholesterol: levels < 40 mg/dL (men) or < 50 mg/dL (women).

MetS status was analyzed as a binary variable (present vs. absent).

#### Covariates

Covariates were chosen based on existing epidemiological evidence concerning stroke risk and their potential to confound the MetS–stroke relationship [1]. The following were included:

- Age group: 19–49, 50–69, or ≥70 years;
- Sex: male or female;
- Educational level: ≤elementary school, middle school, high school, or ≥college;
- Family history of stroke: paternal or maternal history of stroke (yes/no);
- Economic activity status: active or inactive;
- SRH: assessed on a five-point scale (very good, good, fair, poor, and very poor).

### Statistical analysis

All analyses were performed using SPSS Statistics version 25.0 (IBM Corp., Armonk, NY, USA) via the complex samples module. The integrated three-year sampling weight (wt_itvex/3), stratification variable (kstrata), and primary sampling units (psu) were specified throughout the analysis to accommodate the complex survey design of KNHANES.

The baseline characteristics of the study participants according to stroke status are summarized in Table 1. Differences between groups were assessed using complex-sample linear regression for continuous variables and Rao–Scott adjusted chi-square tests for categorical variables.

**Table 1.**
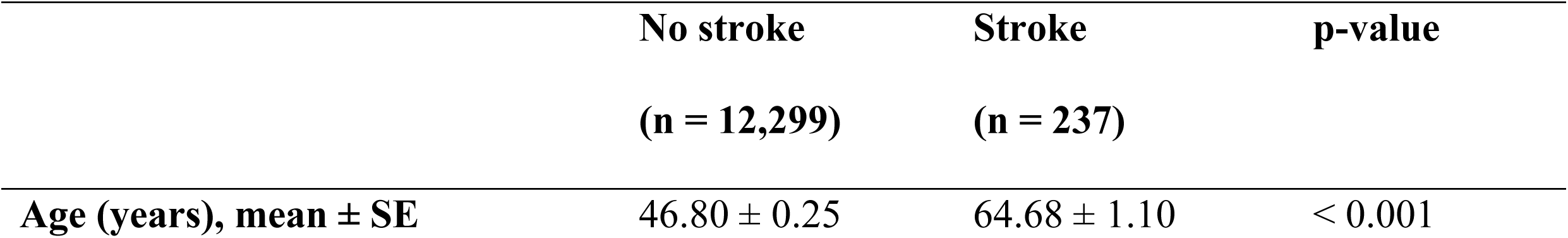
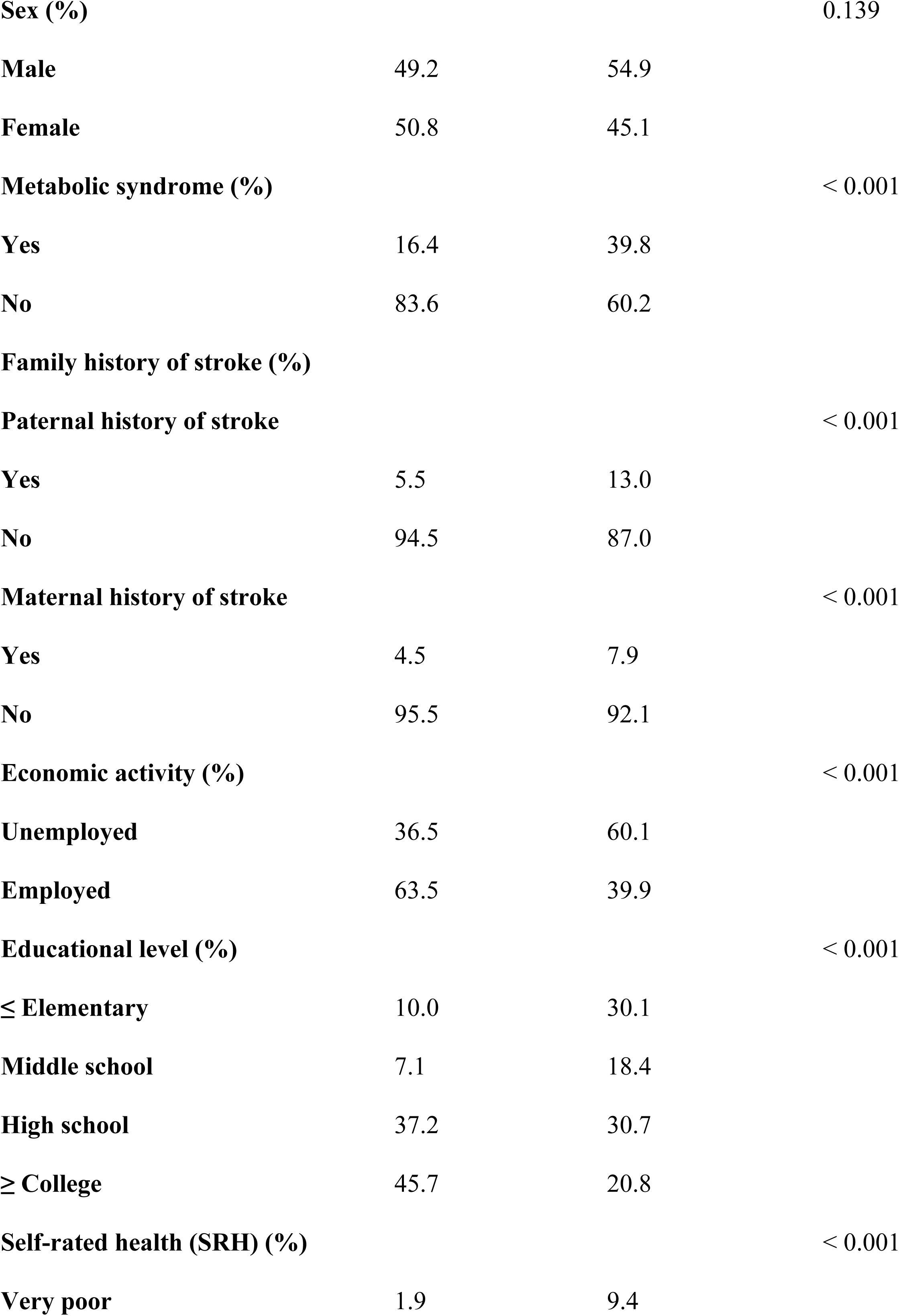
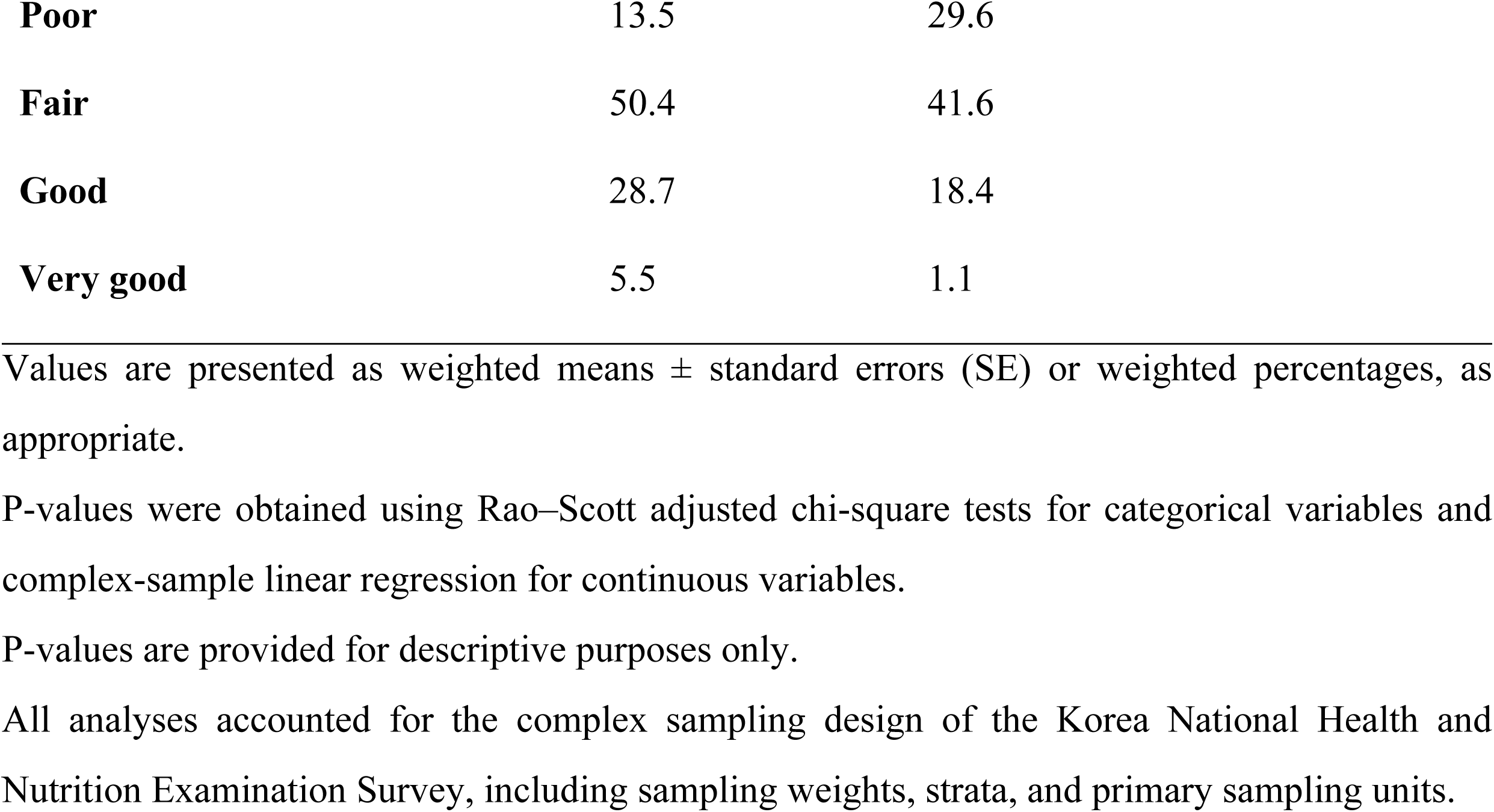
Baseline characteristics of study participants according to stroke status.

|  | No stroke<br>(n = 12,299) | Stroke<br>(n = 237) | p-value |
| --- | --- | --- | --- |
| Age (years), mean ± SE | 46.80 ± 0.25 | 64.68 ± 1.10 | < 0.001 |
| **Sex (%)** |  |  | 0.139 |
| **Male** | 49.2 | 54.9 |  |
| **Female** | 50.8 | 45.1 |  |
| **Metabolic syndrome (%)** |  |  | < 0.001 |
| **Yes** | 16.4 | 39.8 |  |
| **No** | 83.6 | 60.2 |  |
| **Family history of stroke (%)** |  |  |  |
| **Paternal history of stroke** |  |  | < 0.001 |
| **Yes** | 5.5 | 13.0 |  |
| **No** | 94.5 | 87.0 |  |
| **Maternal history of stroke** |  |  | < 0.001 |
| **Yes** | 4.5 | 7.9 |  |
| **No** | 95.5 | 92.1 |  |
| **Economic activity (%)** |  |  | < 0.001 |
| **Unemployed** | 36.5 | 60.1 |  |
| **Employed** | 63.5 | 39.9 |  |
| **Educational level (%)** |  |  | < 0.001 |
| **≤ Elementary** | 10.0 | 30.1 |  |
| **Middle school** | 7.1 | 18.4 |  |
| **High school** | 37.2 | 30.7 |  |
| **≥ College** | 45.7 | 20.8 |  |
| **Self-rated health (SRH) (%)** |  |  | < 0.001 |
| **Very poor** | 1.9 | 9.4 |  |

|  |  |  |
| --- | --- | --- |
| **Poor** | 13.5 | 29.6 |
| **Fair** | 50.4 | 41.6 |
| **Good** | 28.7 | 18.4 |
| **Very good** | 5.5 | 1.1 |

Values are presented as weighted means ± standard errors (SE) or weighted percentages, as appropriate.
P-values were obtained using Rao–Scott adjusted chi-square tests for categorical variables and complex-sample linear regression for continuous variables.
P-values are provided for descriptive purposes only.
All analyses accounted for the complex sampling design of the Korea National Health and Nutrition Examination Survey, including sampling weights, strata, and primary sampling units.

The relationship between MetS and physician-diagnosed stroke was modelled using complex-sample logistic regression under a sequential adjustment scheme (Table 2).

- Model 1: crude (unadjusted) model;
- Model 2: adjusted for age group and sex;
- Model 3: additionally adjusted for educational level and family history of stroke;
- Model 4: further adjusted for economic activity status and SRH.

Estimates were reported as odds ratios (ORs) with 95% confidence intervals (CIs), and significance was defined as a two-sided p-value of <0.05. To convey the explanatory contribution of each successive model, the Nagelkerke R² was reported alongside each OR and its 95% CI for every stage of adjustment.

**Table 2.**
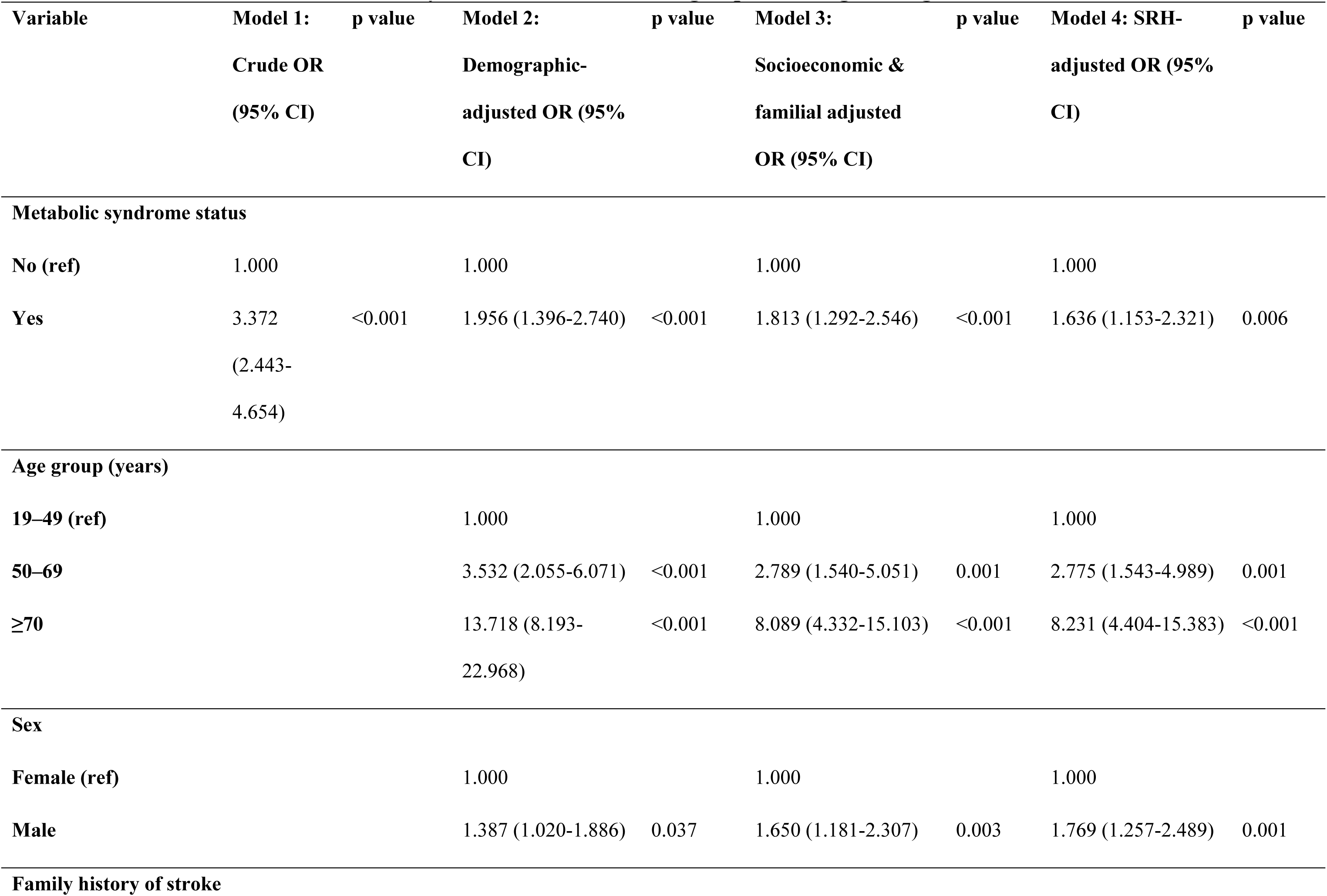
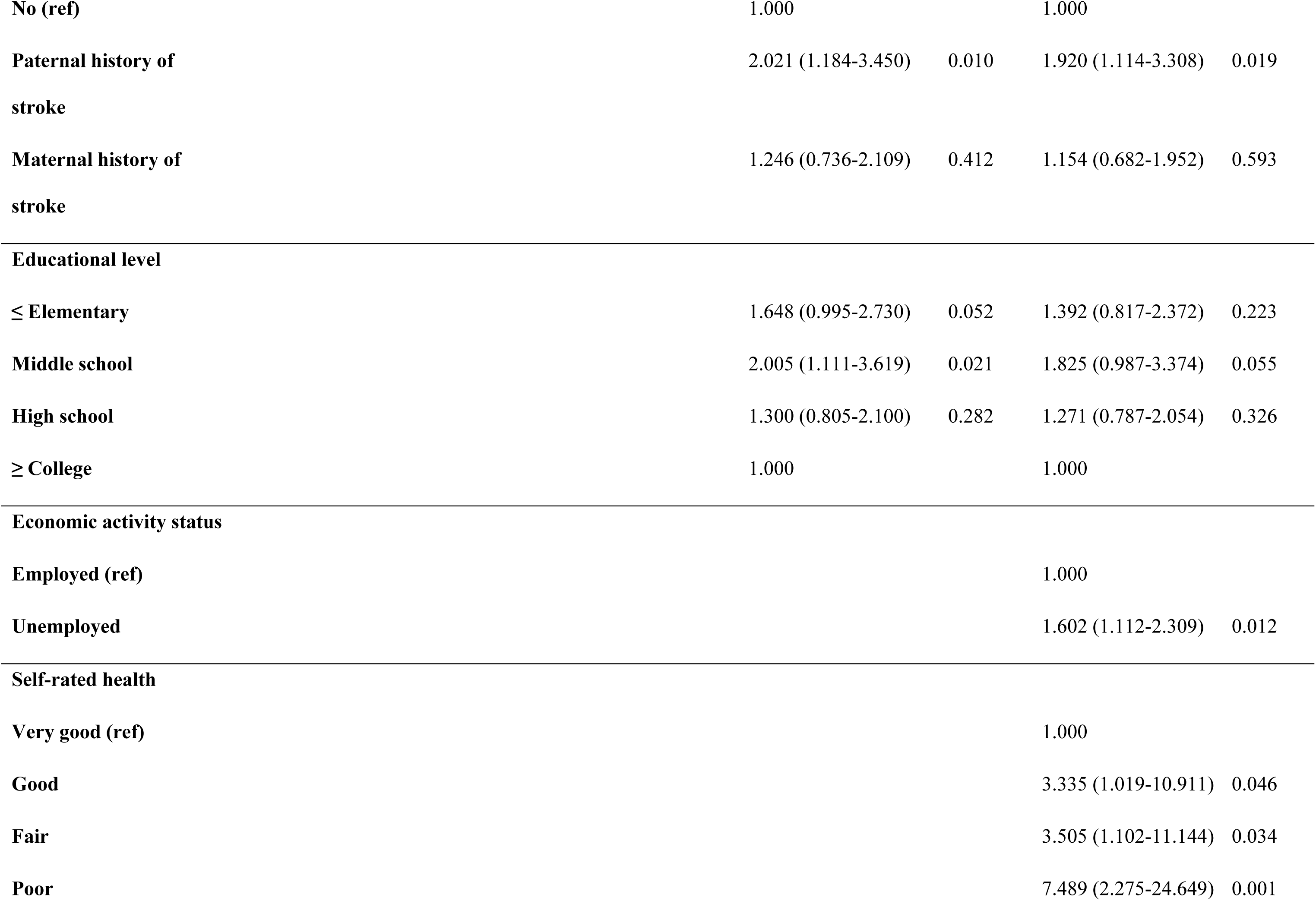
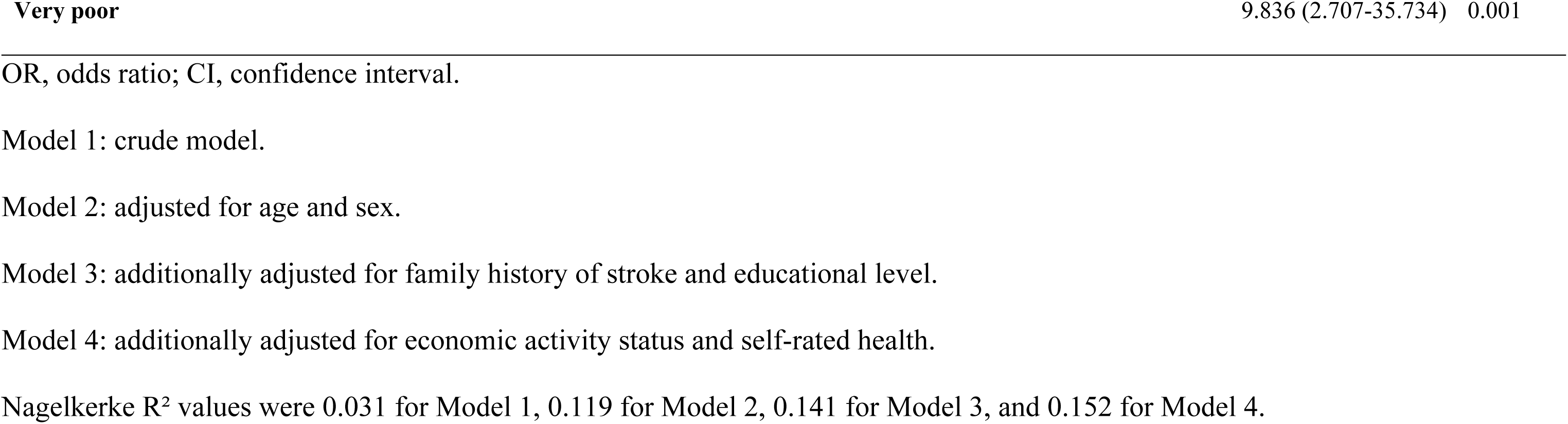
Association between metabolic syndrome and stroke using sequential logistic regression models.

As the KNHANES is a fixed, pre-existing national survey, the sample size was not determined by *a priori* calculation. Therefore, we conducted a *post hoc* evaluation of statistical power for the primary association. With 12,536 weighted observations, 237 stroke cases, and the observed exposure prevalence, the analysis provided well in excess of the 0.80 threshold of the statistical power required to detect the fully adjusted effect size (OR = 1.636) at a two-sided α of 0.05, confirming that the study was adequately powered for the principal comparison.

Covariate selection was guided by prior knowledge of the determinants of MetS and stroke. This approach was consistent with the logic of a directed acyclic graph (DAG), in which age, sex, educational level, family history, economic activity status, and SRH acted as common antecedents of exposure and outcome. A formal DAG was not pre-specified; rather, a sequential adjustment strategy was used to explicitly clarify how the estimated association responded as these conceptually grouped confounders were introduced.

Stratified analyses across the three consolidated SRH categories were conducted to examine whether the MetS–stroke association varied according to the perceived health level (Table 3). Within each SRH stratum, the association was estimated using complex-sample logistic regression adjusted for age, sex, educational level, family history of stroke, and economic activity status. Thus, Model 4’s covariate set omitted SRH, since that variable defined strata. These analyses were exploratory, and no formal interaction tests were performed.

**Table 3.** Stratified analysis by SRH.

| SRH strata | OR (95% CI) | p value |
| --- | --- | --- |
| **Poor** | 1.74 (1.03–2.94) | 0.039 |
| **Fair** | 1.70 (1.05–2.77) | 0.032 |
| **Good** | 1.62 (0.68–3.87) | 0.274 |

Values are presented as odds ratios (ORs) with 95% confidence intervals (CIs).

All models were adjusted for age group, sex, educational level, family history of stroke, and economic activity status.

Self-rated health (SRH) was categorized into poor, fair, and good.

Stratified analyses were conducted to explore potential effect modification by self-rated health.

P-values are provided for descriptive purposes only.

### Ethical considerations

The KNHANES releases publicly available datasets in a fully anonymized form. The dataset analyzed in this study was accessed for research purposes in October 2025. The authors had no access to any information that could identify individual participants during or after data collection. Accordingly, the Institutional Review Board of Yonsei University Mirae Campus approved an exemption from review for this secondary analysis (approval no.: 1041849-202511-BM-244-01), and informed consent was not required.

## Results

### Participant characteristics

Table 1 presents the baseline characteristics of the participants according to stroke status. In total, 237 (1.9%) participants reported a history of physician-diagnosed stroke, whereas 12,299 (98.1%) did not. The participants with stroke were considerably older than those without, with weighted mean ages of 64.68 years (standard error, 1.10) versus 46.80 years (standard error, 0.25).

The weighted prevalence of MetS was 16.8% in the overall cohort. MetS was significantly more common among participants with stroke than among those without stroke (39.8% vs. 16.4%, p < 0.001). Participants with stroke tended to have completed less education and were more likely to report a family history of stroke. They were more likely to be economically inactive and reported poorer SRH. Crude associations between demographic and socioeconomic covariates and stroke are provided in S1 Table.

### Association between MetS and stroke

The relationship between MetS and physician-diagnosed stroke was evaluated using sequential logistic regression models (Table 2). In the crude model (Model 1), MetS was strongly associated with stroke (OR 3.372, 95% CI 2.443–4.654, p < 0.001). Once age group and sex were considered in Model 2, the association weakened but remained significant (OR 1.956, 95% CI 1.396–2.740, p < 0.001).

Adding educational level and family history of stroke (Model 3) produced a further, more modest reduction in the association (OR 1.813, 95% CI 1.292–2.546, p = 0.001).

The specific contribution of SRH to this attenuation is detailed in S2 Table.

### Covariates in the fully adjusted model

Several covariates were independently associated with stroke in our fully adjusted Model 4. Older age was strongly associated with stroke. Compared with those aged 19–49 years, our participants aged 50–69 years had an OR of 2.775 (95% CI 1.543–4.989, p = 0.001) for having had a stroke, whereas those aged ≥70 years had an OR of 8.231 (95% CI 4.404–15.383, p < 0.001). Male sex was independently associated with a higher risk of stroke (OR 1.769, 95% CI 1.257–2.489, p = 0.001).

A paternal history of stroke remained significantly associated with stroke after adjustment, whereas a maternal history did not reach significance in the fully adjusted model. Economic inactivity was associated with a higher odds of stroke (OR 1.602, 95% CI 1.112–2.309, p = 0.012). SRH showed a graded relationship with the risk of stroke. Compared to the participants who reported excellent SRH, those who reported progressively worsening health had higher odds of having a physician-diagnosed stroke.

### Stratified analysis by self-rated health status

Our stratified analyses of the MetS–stroke association across the SRH categories are shown in Table 3. Among the participants who reported poor SRH, MetS was associated with significantly higher odds of stroke (OR 1.74, 95% CI 1.03–2.94, p = 0.039). A comparable association was observed in the fair SRH group (OR 1.70, 95% CI 1.05–2.77, p = 0.032). Conversely, among those who reported good SRH, this association did not reach significance (OR 1.62, 95% CI 0.68–3.87, p = 0.274).

## Discussion

In this nationally representative sample of Korean adults from the KNHANES VIII (2019–2021) dataset, we examined the relationship between MetS and physician-diagnosed stroke, progressively accounting for demographic, socioeconomic, and SRH factors. The initial association was strong (OR 3.37) but decreased substantially as more covariates were added before settling at a fully adjusted OR of 1.64, representing an approximately 51% reduction in effect size. The substantial age gap between the stroke and non-stroke groups in this study (mean ages 64.7 and 46.8 years, respectively) suggests that demographic structure and age explain much of the crude estimate. This attenuation likely reflects the role of these variables as confounders, which represent common causes of both MetS and stroke rather than as mediators along a causal pathway. The sequential approach adopted in this study was intended to expose the confounding structure behind the observed association rather than partition a causal effect. Despite this attenuation, the association persisted in the fully adjusted model, implying that the clustering of metabolic risk maintained an independent relationship with stroke even after accounting for these broader contextual determinants [15,19].

The steepest attenuation followed our adjustments for age and sex, indicating that the demographic structure carried a large share of the crude MetS–stroke association. This is precisely the pattern expected when these variables operate as confounders. Age is a well-established determinant of metabolic disturbance and cerebrovascular disease, and its uneven distribution across MetS categories may spuriously inflate crude estimates. In our data, older age groups showed a strong independent association with stroke, with the highest odds among those aged ≥70 years. These observations reinforce the importance of adjusting for demographics when interpreting cardiometabolic risk associations in population-based studies. The link between metabolic risk clustering and stroke may be overstated if age distribution is not sufficiently controlled for, not because age transmits the biological effect of MetS, but because it confounds the observed relationship [20].

Further adjustments for educational level and family history of stroke yielded a further reduction, indicating that these factors were confounders. Educational attainment is a widely used proxy for socioeconomic status and indicates long-term exposure to structural health determinants, such as health literacy, occupational circumstances, and access to preventive services [21,22]. Individuals with lower educational attainment may have antecedent conditions that independently increase the likelihood of metabolic dysregulation and stroke, generating confounding factors rather than reflecting the downstream effect of MetS. Similarly, a family history of stroke encompasses a genetic predisposition, together with shared lifestyle and environmental exposures that may elevate the risk for both conditions concurrently [23]. In our analysis, a paternal history of stroke was significantly associated with stroke in the fully adjusted model, indicating that familial susceptibility may persist even after accounting for measured metabolic and socioeconomic factors.

The inclusion of SRH was interpreted as an additional confounding adjustment rather than evidence of mediation. Although it is biologically plausible that MetS may lower SRH over time, this hypothesis could not be tested within the cross-sectional framework of this study. This study viewed SRH more conservatively as a marker of broader health vulnerability—spanning comorbidity burden, functional limitation, and psychological state—which predispose individuals to metabolic disturbances and adverse cerebrovascular outcomes, in line with a confounding rather than a mediating role [24].

With this interpretive caveat in mind, SRH displayed a strong graded relationship with stroke risk in our fully adjusted model, with the odds of stroke rising steadily as perceived self-health worsened. This agrees with the previous literature, wherein SRH is a robust predictor of cardiovascular morbidity and mortality and captures certain aspects of health that clinical biomarkers alone do not fully convey [25,26]. Economic inactivity was independently associated with stroke after full model adjustment, although this finding should be interpreted with cautious, given the cross-sectional design of the study. In population-based surveys, economic inactivity may reflect retirement, disability, or health-related withdrawal from work. Therefore, its association with stroke may be related to reverse-causation rather than confounding factors alone [27,28].

Our analyses, stratified by SRH, provided descriptive insights into the contextual structure of the MetS–stroke association. MetS was significantly associated with stroke in participants who reported fair or poor SRH but not in those who reported good health. These results are exploratory and should not be construed as evidence of causal effect modification without further studies. Several explanations are plausible: those who rate their health as poor may have greater multimorbidity or cumulative physiological burden, potentially magnifying the clinical relevance of metabolic risk clustering [25]. Conversely, those who reported good health may have had milder metabolic abnormalities or better-controlled risk factors that weakened the observable association [26]. Regardless, the number of patients with stroke within each stratum was relatively small, and no formal interaction test was conducted.

From a biological standpoint, the residual MetS–stroke association that remained after our comprehensive confounder adjustments was consistent with the recognized pathophysiological pathways linking metabolic dysregulation and cerebrovascular disease. MetS comprises a group of abnormalities, including central obesity, insulin resistance, dyslipidemia, and hypertension, which together foster endothelial dysfunction, chronic inflammation, and accelerated atherosclerosis [29]. These processes can increase the susceptibility of a patient to thrombotic and ischemic events, thereby contributing to strokes through pathways that are plausibly independent of the demographic and socioeconomic confounders we adjusted for in this study [30]. Our findings align with recent large-scale evidence. An independent MetS–stroke association has been reported in the UK Biobank [31], a graded relationship between greater MetS severity and stroke risk has been described in middle-aged and older Chinese adults [32], and MetS was shown to mediate part of the association between adiposity indices and incident stroke in the China Health and Retirement Longitudinal Study cohort [33]. The persistence of a significant association after full adjustment thus supports the biological relevance of MetS as a risk marker, although the present study’s design precludes conclusions regarding causality.

These findings have implications for clinical practice and public health. First, the substantial attenuation observed after demographic adjustment underscores the need to rigorously control for population age structure when interpreting cardiometabolic risk associations at the population level, as failing to do so may greatly overstate the independent contribution of MetS. Second, the significant association in the fully adjusted model supports the continued usefulness of MetS as a practical flag for identifying individuals with multiple cardiometabolic risk factors who may benefit from integrated preventive care [15,19]. Third, the strong independent association between SRH and stroke, regardless of its precise role as a confounder or potential mediator, suggests that a simple subjective health assessment may help identify individuals at an elevated risk of stroke who merit closer clinical attention [25].

This study had several notable strengths. It used the KNHANES VIII, a large, nationally representative dataset with standardized measurements and rigorous probability-based sampling. Applying complex survey weights ensured that our findings were likely applicable to the broader Korean adult population rather than to our cohort exclusively. Sequential modelling was used to elucidate how demographic, socioeconomic, and subjective health factors shaped the observed association. The explicit treatment of the confounding-versus-mediation distinction added a measure of methodological clarity that is often missing from comparable observational work.

However, certain key limitations merit acknowledgment. First, its cross-sectional design inherently precludes conclusions regarding causality [34]. The direction and timing of the relationships among MetS, contextual factors, and stroke cannot be established from a single assessment, and a history of stroke may alter metabolic profiles through related medication, lifestyle changes, or reduced mobility, raising the possibility of reverse causation [34]. Second, the stepwise attenuation of the OR across our models, although informative regarding the confounding structure, was not a formal mediation analysis and should not be regarded as a measure of mediated effects. Third, stroke was based on self-reported physician diagnoses, which may be vulnerable to recall bias or differential misclassification across the MetS categories. Fourth, residual confounding may have remained despite our adjustments for several key covariates, as unmeasured influences (e.g., dietary patterns, physical activity levels, smoking statuses, alcohol uses, medications, and detailed comorbidities) were not fully captured in this study.

## Conclusions

In this nationally representative study of Korean adults, MetS was significantly associated with physician-diagnosed stroke. However, the strength of this association was substantially attenuated when demographic characteristics, socioeconomic factors, and SRH were accounted for, with > 50% of the crude association being due to these contextual determinants. These results indicate that the MetS–stroke relationship arises from a combination of biological mechanisms and broader health-related characteristics within specific populations rather than from an isolated metabolic pathway. In particular, self-rated health may reflect both metabolic burden and overall functional health. Further longitudinal studies are warranted to clarify the related temporal dynamics and delineate the biological and contextual contributions to stroke risk.

## Data Availability

All data analyzed in this study are publicly available from the Korea National Health and Nutrition Examination Survey (KNHANES), conducted by the Korea Disease Control and Prevention Agency (KDCA). The KNHANES VIII (2019–2021) datasets used in this study can be downloaded free of charge from the official KNHANES repository at https://knhanes.kdca.go.kr after a brief, free researcher registration required by the data custodian. No new data were generated by the authors during this study only secondary analysis was performed

## Acknowledgments

The authors have no additional acknowledgments to declare.

## Supporting Information

**S1 Table.** Crude associations of demographic, socioeconomic, and health-related factors with stroke

**S2 Table.** Change in association between metabolic syndrome and stroke after additional adjustment for self-rated health

